# Conspiratorial thinking in a 50-state survey of American adults

**DOI:** 10.1101/2024.09.12.24313575

**Authors:** Roy H. Perlis, Ata Uslu, Sergio A. Barroilhet, Paul A. Vohringer, Mauricio Santillana, Matthew A. Baum, James N. Druckman, Katherine Ognyanova, David Lazer

## Abstract

**Background:** While the NIMH Research Domain Criteria framework stresses understanding how neuropsychiatric phenotypes vary across populations, little is known outside of small clinical cohorts about conspiratorial thoughts as an aspect of cognition.

**Methods:** We conducted a 50-state non-probability internet survey conducted in 6 waves between October 6, 2022 and January 29, 2024, with respondents age 18 and older. Respondents completed the American Conspiratorial Thinking Scale (ACTS) and the 9-item Patient Health Questionnaire (PHQ-9). Survey-weighted regression models were used to examine sociodemographic and clinical associations with ACTS score, and associations with vaccination status.

**Results:** Across the 6 survey waves, there were 123,781 unique individuals. After reweighting, a total of 78.6% of respondents somewhat or strongly agreed with at least one conspiratorial idea; 19.0% agreed with all four of them. More conspiratorial thoughts were reported among those age 25 - 54, males, individuals who finished high school but did not start or complete college, those with household income between $25,000 and $50,000 per year, and those who reside in rural areas, as well as those with greater levels of depressive symptoms. Endorsing more conspiratorial thoughts was associated with a significantly lower likelihood of being vaccinated against COVID-19.

**Discussion:** A substantial proportion of US adults endorsed at least some conspiratorial thinking, which varied widely across population subgroups. The extent of correlation with non-vaccination suggests the importance of considering such thinking in designing public health strategies.

## Introduction

Paranoid experiences are a complex phenomenon that lie on a continuum between normality and psychopathology, an illustration of the continuity emphasized by the NIMH Research Domain Criteria framework.^1,2^ These experiences can manifest as varying degrees of distrust and suspicion towards others, ranging from subtle concerns about others’ intentions to absolute certainty in false beliefs of harm. This spectrum includes phenomena such as feeling watched, spied upon, plotted against, oppressed, or controlled. Conspiratorial thinking falls within this continuum and refers to beliefs that are out of the mainstream or do not align with reality.^3^ Such thoughts may represent a response to uncertainty, addressing feelings of insecurity and vulnerability.^4^

A 2021 U.S. survey found that between 9 and 12% of adults endorsed conspiratorial beliefs (e.g., that moon landings were faked, or vaccination implants microchips); another 10% were unsure about these ideas.^5^ While such thoughts have received more attention in the context of the pandemic, a prior report suggested that belief in conspiracies has remained relatively stable over time.^6^ While not inherently pathological,^7^ at the extreme conspiracy beliefs can profoundly impact functioning.^8,9^

Dimensions of functioning have nearly always been investigated in clinically defined subsets, as they are challenging to characterize at population scale. Traditional epidemiologic studies are extremely costly to conduct and have most often focused on standard diagnostic categories. However, we have previously shown that an alternative approach, using large-scale nonprobability internet sampling with quotas and rigorous design, can yield valid estimates of population characteristics.^10^

Here, we drew on a 50-state U.S. survey of adults to examine variation in conspiracy-mindedness, using a measure focused on beliefs about the U.S. We examined the extent to which conspiracy-minded beliefs vary across adults, and in particular whether they correlate with depressive symptoms. We then sought to understand their association with public health behaviors as exemplified by vaccination, hypothesizing that conspiratorial thought would predispose to avoiding government-advocated health measures.

## Methods

### Study Design

We conducted a nonprobability ^11^ web-based survey via a commercial survey panel aggregator, Pure Spectrum, as part of an academic consortium called the COVID States Project; the 6 survey waves reported here (Waves #25 - 30) were conducted between October 6, 2022 and January 29, 2024. The survey used state-by-state quotas for age, gender, and race and ethnicity to yield representative samples. Eligible participants were 18 and older and resided in the United States; they could opt in to a general survey of opinions (rather than a survey of beliefs on a particular topic, for example) in return for compensation that varied by the panel. Each panel provided an incentive for respondents that could vary depending on survey length and panelist profile; these incentives could include cash payments, airline miles, gift cards, redeemable points (for example, for mobile games), entrance into a sweepstakes, and vouchers. All participants consented online to participation before answering survey questions. The survey protocol was evaluated and considered to be exempt by the Harvard University Institutional Review Board. We present survey results in accordance with AAPOR guidelines. ^12^

### Measures

Conspiratorial thinking was assessed by the American Conspiratorial Thinking Scale (ACTS).^13^ This measure asks, “How much do you agree or disagree with the following statements?” followed by 4 questions: “Even though we live in a democracy, a few people will always run things anyway,” “The people who really ’run’ the country are not known to the voters,” “Big events like wars, the current recession, and the outcomes of elections are controlled by small groups of people who are working in secret against the rest of us,” and “Much of our lives are being controlled by plots hatched in secret places.” Each question is answered on a 1-5 scale: “strongly disagree” (1), “somewhat disagree (2), “neither agree nor disagree” (3), “somewhat agree” (4), and “strongly agree” (5). To simplify analysis of this ordinal scale, we dichotomized responses to identify those who agree (4 or 5) or do not agree (1-3) with each statement.

Depressive symptoms severity was assessed with the 9-item Patient Health Questionnaire (PHQ-9),^14,15^ reflecting diagnostic criteria for major depressive disorder in the DSM-5. Participants were asked to describe their social network in terms of number of individuals who provide social support in each of 4 domains (“Now please think of your complete social circle of family, friends, neighbors, and other acquaintances. Approximately how many of them could you count on for the following things? – To…”) followed by medical care, financial support, emotional support, and help with employment.^16^ They were also asked to identify the number of individuals other than them who reside in their home.

We also collected sociodemographic features, to confirm representativeness of the US population and facilitate survey weighting and subgroup analyses. They were asked to identify race and ethnicity from a list including African American or Black, Asian American, Hispanic, Native American, Pacific Islander, white, or Other, and could provide a free text self-description. To facilitate inclusion of smaller groups, we collapsed Native America, Asian-Pacific Islander, and Other into a single category for analysis, and dichotomized employment status to “working full-time” (yes vs. all others).

### Statistical Analysis

We first examined associations between individual sociodemographic features and number of conspiracy items endorsed, using linear regression. As sensitivity analysis, repeating analyses using ordinal logistic regression as implemented in the *polr* package in R did not yield meaningfully different estimates of effect. In these and all subsequent regression models, additional covariates included age category (to allow for nonlinear effects), gender, education (categorized as graduate, undergraduate, some college, high school graduate, some high school or less), annual household income (categorized as <$25k, $25-<$50k, $50k-<$100k, >$100k), race and ethnicity, and rural, suburban, or urban setting. We then examined the additional association with social supports in 4 categories, and number of individuals at home. These analyses considered numbers to be categories, to allow for non-linear associations, and *a priori* truncated count of supports in each category at 5.

Survey weights were applied to estimate national distributions, using the R survey package (version 4.2-1).^17^ We applied interlocking national weights for age at survey completion, sex, and race and ethnicity, as well as education and region, using 2019 US Census American Community Survey data,^18^ a standard approach for nonprobability samples.^19^ (For generation of choropleths reflecting state-level prevalence, we used corresponding state values from the 2019 Census.)

As individuals could respond to more than one survey wave, we selected the initial (index) response for cross-sectional analyses. In prior analyses, sensitivity analysis such as random selection of a response, or considering multiple responses as clustered within an individual, did not yield meaningfully different results.^20^ To examine change over time, we secondarily analyzed the subset of individuals who completed more than one survey, identifying as baseline the initial survey and follow-up the next survey completed. These analyses used R 4.3.2 ^21^, and considered p <.05 to represent statistical significance.

## Results

Across the 6 survey waves, there were 123,781 unique individuals. Mean age was 46.6 (SD 17.1) years; 74,570 (60.2%) identified as women, 47,791 (38.6%) as men, and 1,420 (1.1%) as nonbinary. A total of 4,235 (3.4%) identified as Asian, 16,306 (13.2%) as Black or African American, 13,501 (10.9%) as Hispanic, 1,589 (1.3%) as Native American, 1,364 (1.1%) as Pacific Islander, 1,968 (1.6%) as another race or ethnicity, and 84,818 (68.5%) as white. Additional characteristics of the cohort are summarized in Table 1.

**Table 1.** Characteristics of survey respondents included in analyses of conspiratorial thinking, October 2022-January 2024.

|  | <4 ideas endorsed<br>(N=100951) | All 4 ideas endorsed<br>(N=22830) | Total<br>(N=123781) |
| --- | --- | --- | --- |
| **Age in years (SD)** | 46.8 (17.2) | 45.9 (16.5) | 46.6 (17.1) |
| **Gender** |  |  |  |
| Female | 62552 (62.0%) | 12018 (52.6%) | 74570 (60.2%) |
| Male | 37170 (36.8%) | 10621 (46.5%) | 47791 (38.6%) |
| Nonbinary | 1229 (1.2%) | 191 (0.8%) | 1420 (1.1%) |
| **Race and Ethnicity** |  |  |  |
| African American | 13240 (13.1%) | 3066 (13.4%) | 16306 (13.2%) |
| Asian American | 3576 (3.5%) | 659 (2.9%) | 4235 (3.4%) |
| Hispanic | 10833 (10.7%) | 2668 (11.7%) | 13501 (10.9%) |
| Native American | 1255 (1.2%) | 334 (1.5%) | 1589 (1.3%) |
| Other (a) | 1573 (1.6%) | 395 (1.7%) | 1968 (1.6%) |
| Pacific Islander | 1069 (1.1%) | 295 (1.3%) | 1364 (1.1%) |
| White | 69405 (68.8%) | 15413 (67.5%) | 84818 (68.5%) |
| **Education** |  |  |  |
| Some High School or Less | 3546 (3.5%) | 781 (3.4%) | 4327 (3.5%) |
| High School Graduate | 22694 (22.5%) | 5768 (25.3%) | 28462 (23.0%) |
| Some College | 25561 (25.3%) | 6071 (26.6%) | 31632 (25.6%) |
| College Degree | 35873 (35.5%) | 7752 (34.0%) | 43625 (35.2%) |
| Graduate Degree | 13277 (13.2%) | 2458 (10.8%) | 15735 (12.7%) |
| **Employment (full-time)** | 39236 (38.9%) | 9714 (42.5%) | 48950 (39.5%) |
| **Income** |  |  |  |
| Under 25K | 22143 (21.9%) | 5263 (23.1%) | 27406 (22.1%) |
| 25k to under 50k | 26467 (26.2%) | 6391 (28.0%) | 32858 (26.5%) |
| 50K to under 100K | 32713 (32.4%) | 7057 (30.9%) | 39770 (32.1%) |
| 100K and over | 19628 (19.4%) | 4119 (18.0%) | 23747 (19.2%) |
| **Urbanicity** |  |  |  |
| Rural | 16983 (16.8%) | 4438 (19.4%) | 21421 (17.3%) |
| Suburban | 58006 (57.5%) | 12406 (54.3%) | 70412 (56.9%) |
| Urban | 25962 (25.7%) | 5986 (26.2%) | 31948 (25.8%) |
| **Conspiracy items** | 1.4 (1.0) | 4.0 (0.0) | 1.9 (1.4) |
| **PHQ9 Total (b)** |  |  |  |
| PHQ9 Total (b) | 6.1 (6.4) | 7.3 (7.3) | 6.4 (6.6) |
| **PHQ9 10 or greater (b)** | 24335 (24.7%) | 6957 (31.2%) | 31292 (25.9%) |
| **Social support (health care) (c)** | 2.5 (1.4) | 2.3 (1.4) | 2.5 (1.4) |
| **Social support (financial) (d)** | 2.1 (1.4) | 2.0 (1.4) | 2.1 (1.4) |
| **Social support (emotional) (e)** | 2.7 (1.4) | 2.5 (1.4) | 2.6 (1.4) |
| **Social support (employment) (f)** | 2.1 (1.6) | 2.0 (1.6) | 2.1 (1.6) |
| **SARS-CoV-2 vaccine** | 77293 (76.6%) | 13909 (60.9%) | 91202 (73.7%) |
| **Influenza vaccine** | 45222 (54.5%) | 8026 (41.9%) | 53248 (52.2%) |
| **More than one survey** | 7513 (7.4%) | 1553 (6.8%) | 9066 (7.3%) |

(a) Other refers to individuals who checked the 'Other race or ethnicity' box from a list of choices.
(b) PHQ-9 was not completed for n=2765 (2260 and 505, respectively)
(c) Social support item 1 was not completed for n=416 (330 and 86, respectively)
(d) Social support item 2 was not completed for n=570 (453 and 117, respectively)
(e) Social support item 3 was not completed for n=740 (585 and 155, respectively)
(f) Social support item 4 was not completed for n=844 (694 and 150, respectively)
(g) Influenza vaccination was not asked for n=21709 (18016 and 3693, respectively)

In all, after reweighting, 78.6% individuals somewhat or strongly agreed with at least one conspiratorial thought; 19.0% agreed with all four of them; Table S1 lists proportion agreeing with each item. Figure 1 illustrates reweighted proportion of individuals endorsing conspiratorial thoughts by state. Among individuals with no evidence of depression by PHQ-9 (i.e., PHQ-9 <5), 75.1% endorsed at least one conspiratorial thought, and 17.3% endorsed all four.

**Figure 1.**
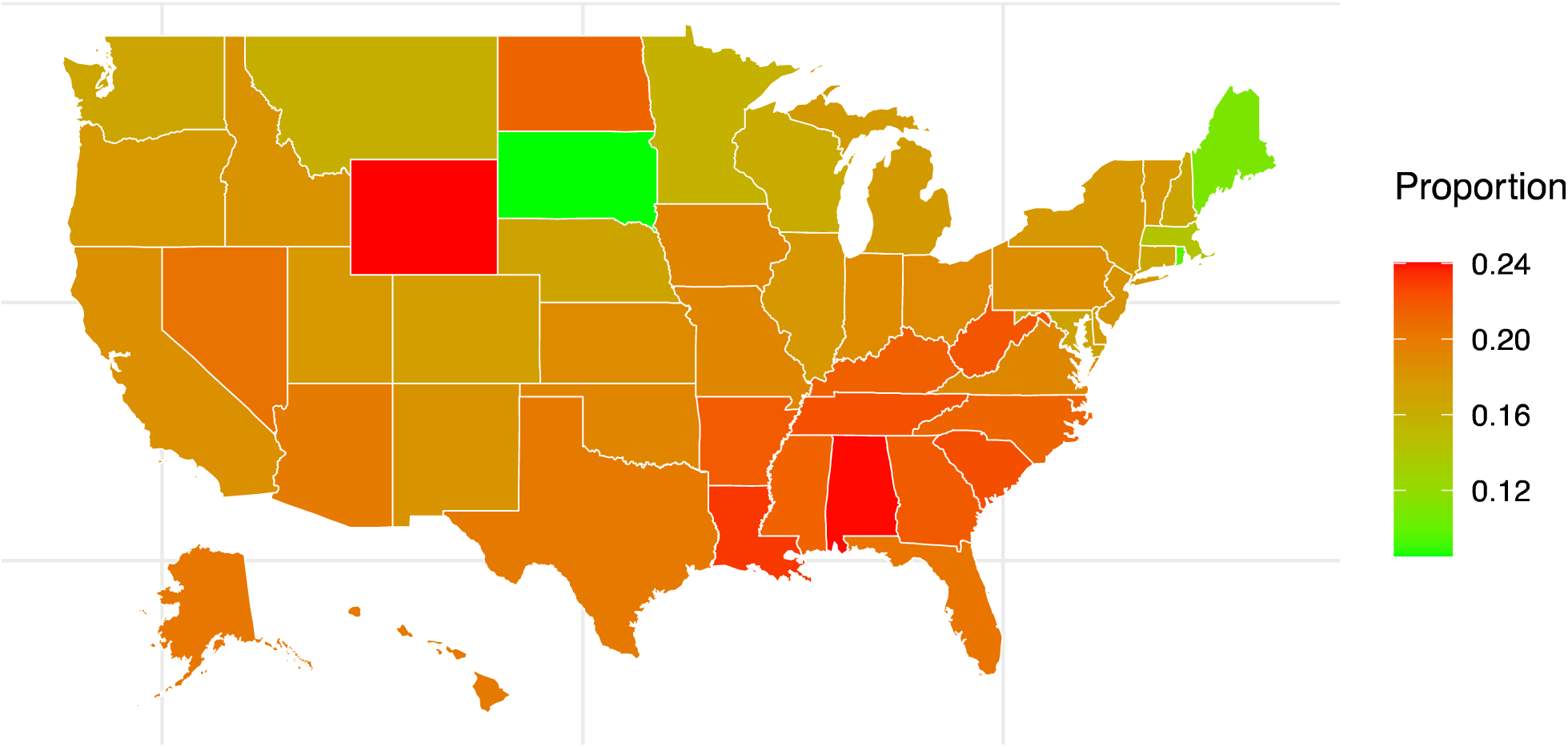
Proportion of US adults endorsing 4 conspiracy items, by state, October 2022-January 2024

In linear regression models, we first examined associations between sociodemographic features and number of conspiracy items endorsed, estimating effects in univariate (Table S2) and fully adjusted models (Figure 2). Subgroups endorsing greater numbers of conspiratorial thoughts included those age 25 - 54, males, individuals who finished high school but did not start or complete college, those with household income between $25,000 and $50,000 per year, and those who reside in rural areas. A greater number of depressive symptoms was also modestly but significantly associated with greater number of conspiratorial thoughts endorsed (coefficient adjusted for sociodemographic features = 0.02, 95% CI 0.02 - 0.02), as was presence of moderate or greater depressive symptoms, coefficient adjusted for sociodemographic features = 0.28, 95% CI 0.26 - 0.30). Incorporating political affiliation did not meaningfully change these associations (Figure S1).

**Figure 2.**
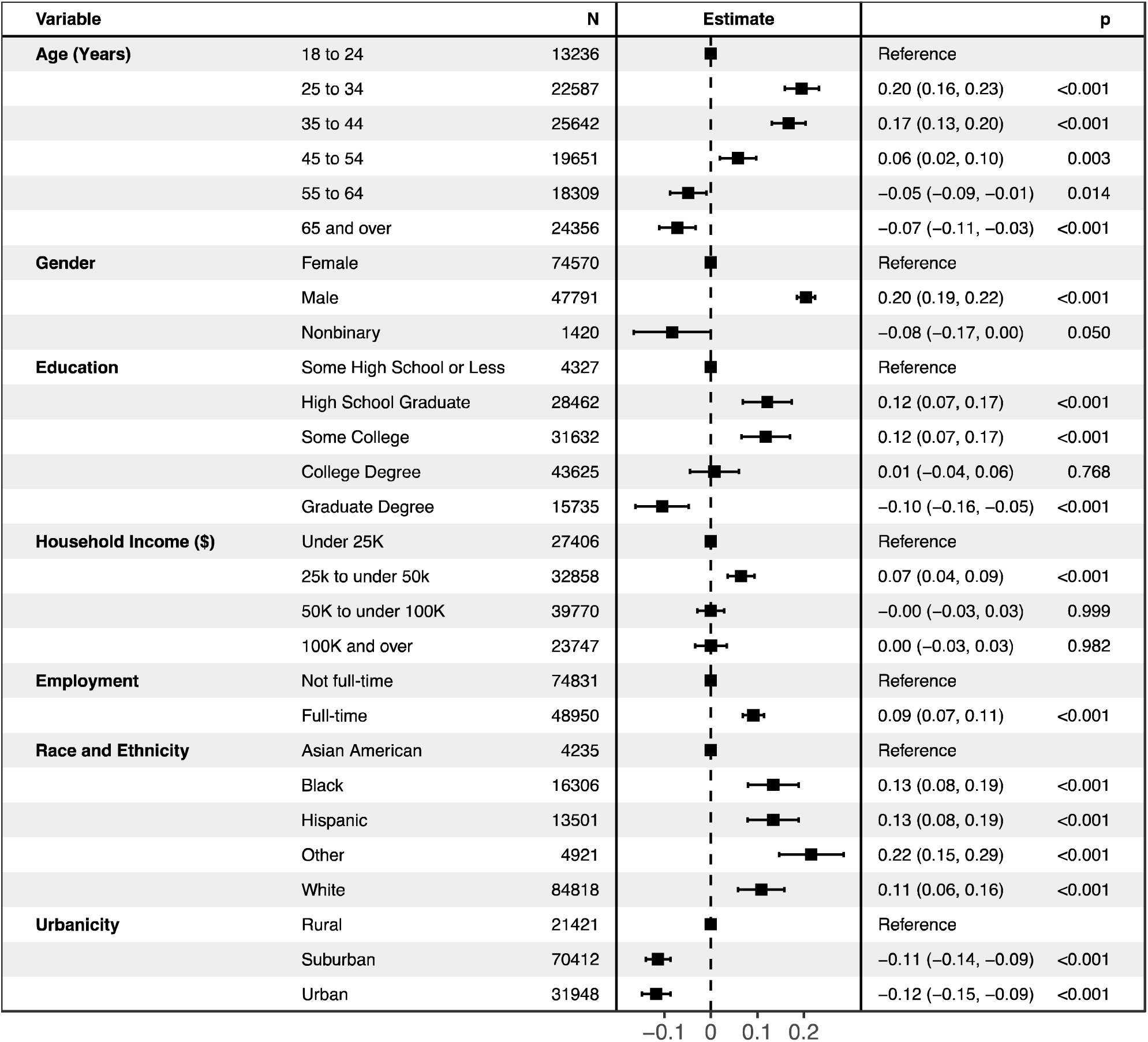
Survey-weighted linear regression model of association between sociodemographic features and number of conspiracy items endorsed

For 8,493 individuals who returned for a subsequent survey, we examined whether depression and conspiracy-mindedness changed in parallel – i.e., whether the changes were correlated. Change in PHQ-9 was significantly associated with change in count of conspiratorial ideas, although these effects were extremely modest (unadjusted coefficient 0.01, 95% CI 0.00 - 0.01; adjusted coefficient 0.01, 95% CI 0.00 - 0.02).

We also examined the association between conspiratorial thoughts and measures of social network size. Figure 3 illustrates these associations for each of the social support measures (supports for health care, financial support, emotional support, and employment support) in survey-weighted regression models adjusted for sociodemographic features. A qualitatively similar pattern emerged across all 4 domains, with endorsement of all 4 conspiratorial thoughts associated with diminished network size. On the other hand, endorsing 1 or 2 such thoughts was significantly associated with a larger network.

**Figure 3.**
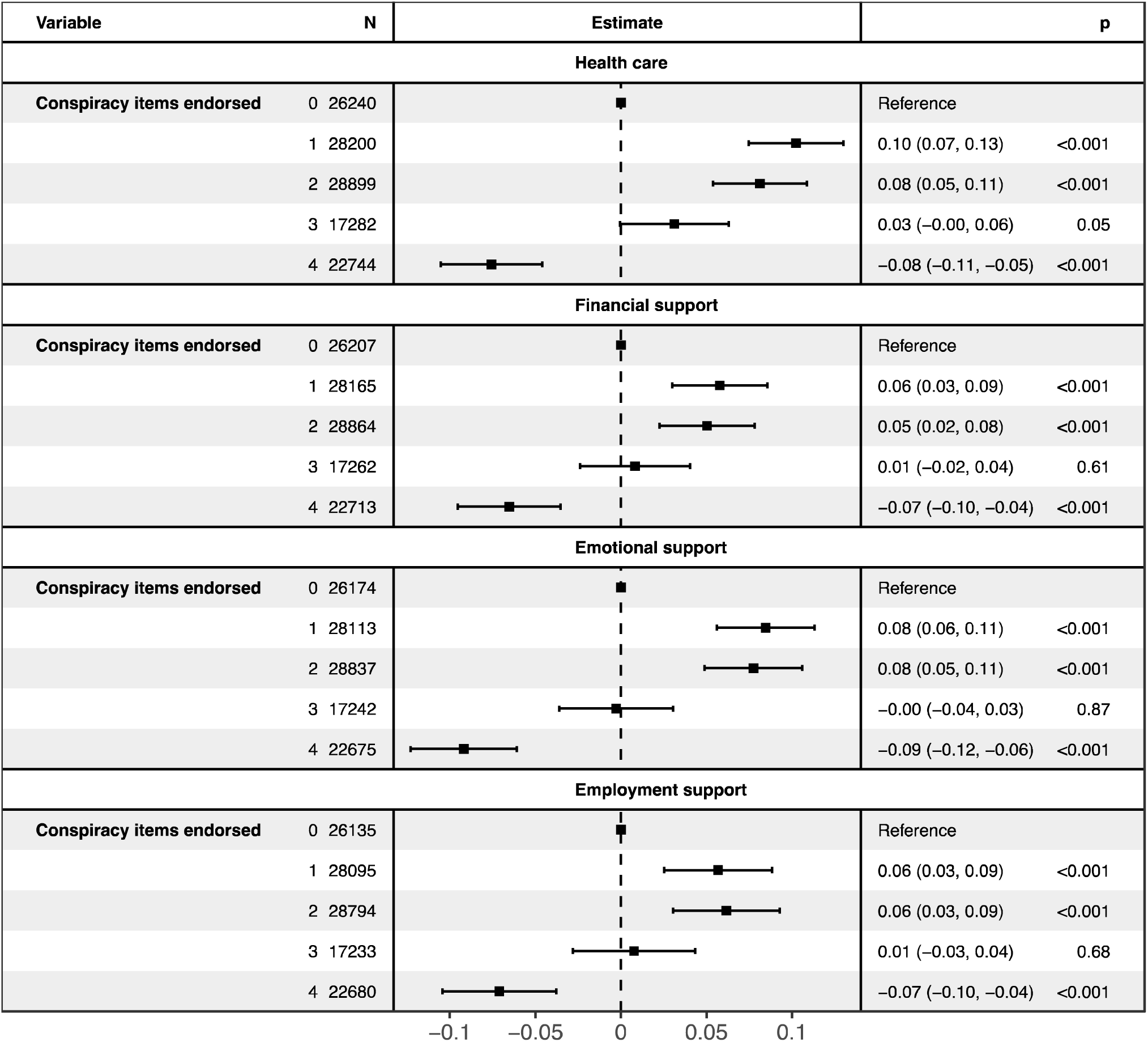
Survey-weighted linear regression models of association between conspiratorial thoughts and individual measures of social network size

Finally, we investigated the association between conspiratorial thoughts and health behavior, focusing on receipt of COVID-19 vaccination and influenza vaccination. In adjusted logistic regression models, a greater number of conspiratorial thoughts endorsed was associated with a lesser likelihood of being vaccinated against COVID-19 (Figure 4); adjusted OR were 0.83 (95% CI 0.79 - 0.88), 0.57 (95% CI 0.54 - 0.60), and 0.43 (95% CI 0.41 - 0.46) among those endorsing 2, 3, or 4 conspiratorial ideas, respectively, compared to 0. Influenza vaccination followed a similar pattern, with adjusted OR of 0.81 (95% CI 0.77 - 0.85), 0.66 (95% CI 0.63 - 0.70), and 0.58 (95% CI 0.55 - 0.61) (Figure S2). Among individuals who returned for a subsequent survey, endorsing a greater number of conspiratorial thoughts was associated with less likelihood of receiving an additional vaccine or booster in the intervening period (Figure S3; adjusted OR 0.72 (95% CI 0.54 - 0.96) for endorsing 4 versus 0 ideas).

**Figure 4.**
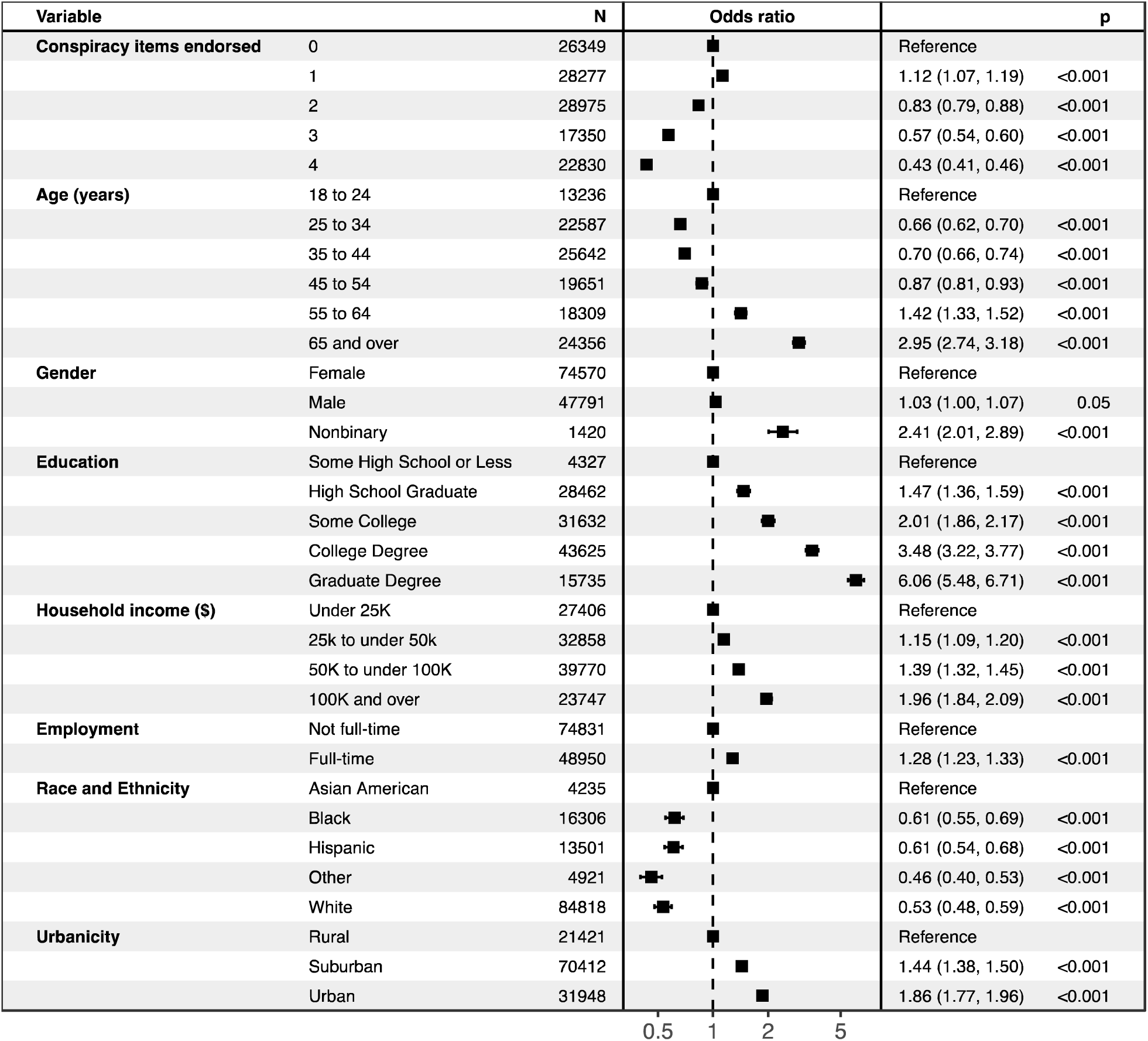
Survey-weighted logistic regression models of association between conspiratorial thoughts and likelihood of being vaccinated against COVID-19

## Discussion

Among a cohort of more than 123,000 US adults, we found that around 3 in 4 respondents endorsed at least one conspiratorial idea, and nearly 1 in 5 endorsed all 4. These ideas were more common among younger individuals, particularly men, as well as those who did not complete college and reside in rural areas. While such ideas were slightly more common among individuals with depressive symptoms, the observed sociodemographic associations were not meaningfully different.

Our results align with a prior investigation suggesting high levels of conspiratorial thinking during the COVID-19 pandemic.^22^ In that study of 2,500 adults across 5 countries, 29% reported conspiratorial ideas related to the pandemic; in the US subsample, male gender and younger age associated with these ideas. An international poll early in the pandemic likewise found a large proportion of individuals endorsing such ideas.^23^

Prior to the pandemic, a UK study ^7^ of paranoia found that around 15 - 20% of individuals endorsed mistrust of others, and up to 10% ideas of reference or persecution. Another pre-pandemic epidemiologic study examined a single item from the National Comorbidity Survey-Replication study, estimating that 27% of U.S. adults endorsed a belief in conspiracy ‘behind many things in the world.’^24^ Notably, that study also found such beliefs to be more common among men and individuals with less education.

The individuals we identified with greatest prevalence of conspiratorial thinking may be those at elevated risk for other factors including early adversity, social isolation and exclusion, lack of social support, lower education, and substance use,^25^ some of which have also been associated with paranoid symptoms.^26^ We cannot directly address other risk factors (e.g., genetic predisposition and temperament), but all of these risks likely interact to contribute to paranoid experiences and conspiratorial thinking. Further underscoring the complexity of these relationships, we identified a nonlinear association between endorsing conspiratorial thoughts and social supports: while endorsing all 4 such thoughts associates with lower levels of support, endorsing 1 or 2 thoughts associates with higher levels of support. Causation, if any, cannot be inferred from our cross-sectional analyses, but broadly speaking our results suggest that presence of conspiratorial thinking in and of itself does not correlate with poorer social functioning.

We also identified a modest association between endorsement of conspiratorial ideas and depressive symptoms. The experience of frank psychosis in more severe mood disorders is well-described, but the phenomenon we characterize is likely substantially more subtle given that the prevalence of psychotic depression has been estimated to be less than 1%.^27^ Individuals with depression may exhibit a range of cognitive deficits ^28^ that may correlate with acceptance of conspiratorial ideas.^29^ In prior work, we showed that depressive symptoms associated with endorsement of misinformation about the COVID-19 pandemic, for example.^30^

Finally, we found that those individuals endorsing greater levels of conspiratorial ideas were less likely to be vaccinated against COVID-19 as well as influenza. Similarly, in the subset of respondents who returned for a second survey, those endorsing conspiratorial ideas initially were also less likely to have received additional COVID-19 vaccination. This result aligns with an array of findings that conspiratorial thinking during the pandemic has been associated with a range of adverse public health outcomes (for a review, see Leonard^31^, which also emphasizes the importance of studying and publicizing this phenomenon). A particular strength of the present study is that it focused on non-health-related conspiratorial thinking, indicating that these thoughts in general relate to health behavior, even when they have nothing to do with health per se.

### Limitations

This study has some limitations. To begin with, we cannot draw conclusions about causation from the associations between conspiratorial thinking and vaccination behaviors. While those behaviors are unlikely to cause individuals to embrace ideas about conspiracies (i.e., reverse causation), some confounding variable could readily contribute to both. Our analysis of panel data at least suggests that the conspiratorial thoughts precede receipt of additional vaccination, and consideration of political affiliation in addition to other sociodemographic features addresses potential confounding. Still, additional longitudinal investigation, and perhaps randomized intervention studies, would be required to better understand causation. Finally, because we employed a nonprobability design as the only feasible way to sample at this scale at low cost, we cannot report response rates. While these designs may not be as robust as probability designs, this survey has been shown to yield valid results in comparison with both administrative data (e.g., firearms) and probability sampling^10,32^. Moreover, recent large probability surveys indicate that a very large proportion of older adults, low-income individuals, and those without college degrees use the internet,^33^ suggesting that the risk of under sampling these groups is not substantial. Likewise, studies of individuals with serious mental illness also indicate high rates of internet usage.^34^

### Conclusion

We found that conspiratorial thinking was common, but differed substantially across the population, exhibiting greatest prevalence among younger individuals, males, those with lower levels of education, those with household income between $25,000 and $50,000, and those residing in more rural areas. While it was significantly associated with depressive symptoms, the magnitude of this association was modest. Respondents who endorsed greater levels of such thinking were less likely to pursue vaccination, underscoring the importance of better understanding these thoughts and how they vary in both health and disease. While these analyses cannot establish causation, strategies to address conspiratorial thoughts may represent an opportunity to improve adherence to some public health initiatives, at least in the United States.

## Acknowledgements

The authors have no acknowledgements to report

## Author Contributions

Roy H. Perlis, MD MSc -- analyzed data, drafted and revised manuscript

Ata Uslu, MS - collected data, revised manuscript

Sergio Barroilhet, MD PhD – revised manuscript

Paul Vohringer, MD MSc MPH – revised manuscript

Mauricio Santillana, PhD – collected data, revised manuscript

James N. Druckman, PhD – collected data, revised manuscript

Matthew A. Baum, PhD – collected data, revised manuscript

Katherine Ognyanova, PhD – collected data, revised manuscript

David Lazer, PhD - collected data, revised manuscript

## Author Approval

All authors have seen and approved the manuscript.

## Data Availability Statement

The survey used for this study is available from the corresponding author for non-commercial use.

