## Supplemental Materials for "Conspiratorial thinking in a 50-state survey of American adults"

**Table S1.** Conspiracy scores, by item

| | <4 endorsed (N=100,951) | All 4 endorsed (N=22,830) | Total (N=123,781) |
|---|---|---|---|
| 1. Even though we live in a democracy, a few people will always run things anyway | | | |
| Strongly disagree | 5,677 (5.6%) | 0 (0.0%) | 5,677 (4.6%) |
| Somewhat disagree | 9,248 (9.2%) | 0 (0.0%) | 9,248 (7.5%) |
| Neither agree nor disagree | 23,622 (23.4%) | 0 (0.0%) | 23,622 (19.1%) |
| Somewhat agree | 44,708 (44.3%) | 9,854 (43.2%) | 54,562 (44.1%) |
| Strongly agree | 17,696 (17.5%) | 12,976 (56.8%) | 30,672 (24.8%) |
| 2. The people who really 'run' the country are not known to the voters | | | |
| Strongly disagree | 10,663 (10.6%) | 0 (0.0%) | 10,663 (8.6%) |
| Somewhat disagree | 15,090 (14.9%) | 0 (0.0%) | 15,090 (12.2%) |
| Neither agree nor disagree | 32,307 (32.0%) | 0 (0.0%) | 32,307 (26.1%) |
| Somewhat agree | 30,652 (30.4%) | 8,956 (39.2%) | 39,608 (32.0%) |
| Strongly agree | 12,239 (12.1%) | 13,874 (60.8%) | 26,113 (21.1%) |
| 3. Big events like wars, the current recession, and the outcomes of elections are controlled by small groups of people who are working in secret against the rest of us | | | |
| Strongly disagree | 21,466 (21.3%) | 0 (0.0%) | 21,466 (17.3%) |
| Somewhat disagree | 20,518 (20.3%) | 0 (0.0%) | 20,518 (16.6%) |
| Neither agree nor disagree | 40,416 (40.0%) | 0 (0.0%) | 40,416 (32.7%) |
| Somewhat agree | 14,416 (14.3%) | 11,520 (50.5%) | 25,936 (21.0%) |
| Strongly agree | 4,135 (4.1%) | 11,310 (49.5%) | 15,445 (12.5%) |
| 4. Much of our lives are being controlled by plots hatched in secret places. | | | |
| Strongly disagree | 29,003 (28.7%) | 0 (0.0%) | 29,003 (23.4%) |
| Somewhat disagree | 20,282 (20.1%) | 0 (0.0%) | 20,282 (16.4%) |
| Neither agree nor disagree | 37,235 (36.9%) | 0 (0.0%) | 37,235 (30.1%) |
| Somewhat agree | 11,338 (11.2%) | 12,508 (54.8%) | 23,846 (19.3%) |
| Strongly agree | 3,093 (3.1%) | 10,322 (45.2%) | 13,415 (10.8%) |

**Table S2.** Univariate associations between individual sociodemographic features and count of conspiracy items endorsed

| Feature | Term | Estimate | Lower CI | Upper CI |
|---|---|---|---|---|
| Age (vs 18-24) | 25-34 | 0.20 | 0.16 | 0.23 |
| | 35-44 | 0.16 | 0.12 | 0.19 |
| | 45-54 | 0.05 | 0.01 | 0.09 |
| | 55-64 | -0.07 | -0.11 | -0.03 |
| | 65+ | -0.12 | -0.15 | -0.08 |
| Gender (vs Female) | Male | 0.21 | 0.19 | 0.23 |
| | Nonbinary | -0.07 | -0.15 | 0.01 |
| Race and Ethnicity (vs Asian) | Black | 0.16 | 0.11 | 0.21 |
| | Hispanic | 0.19 | 0.14 | 0.25 |
| | Other | 0.24 | 0.18 | 0.31 |
| | White | 0.12 | 0.07 | 0.17 |
| Education (vs Not High School) | High school graduate | 0.14 | 0.08 | 0.19 |
| | Some college | 0.13 | 0.08 | 0.18 |
| | College degree | 0.04 | -0.01 | 0.10 |
| | Graduate degree | -0.08 | -0.13 | -0.02 |
| Employment | Full time | 0.16 | 0.14 | 0.18 |
| Income (vs <$25k) | $25k to <$50k | 0.09 | 0.06 | 0.12 |
| | $50k to <$100k | 0.02 | -0.01 | 0.04 |
| | $100k+ | -0.01 | -0.04 | 0.02 |
| Urbanicity (vs Rural) | Suburban | -0.12 | -0.14 | -0.09 |
| | Urban | -0.09 | -0.12 | -0.07 |
| PHQ9 | PHQ9 total | 0.02 | 0.02 | 0.02 |
| | PHQ9 (10+) | 0.28 | 0.26 | 0.30 |

**Figure S1.** Survey-weighted linear regression model of association between sociodemographic features and number of conspiracy items endorsed, including depressive symptoms and political party

| Variable | | N | Estimate | p |
|---|---|---|---|---|
| **Age (years)** | 18 to 24 | 12979 | Reference | |
| | 25 to 34 | 22090 | 0.21 (0.17, 0.24) | <0.001 |
| | 35 to 44 | 25043 | 0.20 (0.16, 0.23) | <0.001 |
| | 45 to 54 | 19105 | 0.08 (0.04, 0.12) | <0.001 |
| | 55 to 64 | 17881 | −0.01 (−0.05, 0.03) | 0.49 |
| | 65 and over | 23685 | −0.01 (−0.05, 0.03) | 0.68 |
| **Gender** | Female | 72751 | Reference | |
| | Male | 46646 | 0.18 (0.17, 0.20) | <0.001 |
| | Nonbinary | 1386 | −0.05 (−0.13, 0.03) | 0.22 |
| **Education** | Some High School or Less | 4200 | Reference | |
| | High School Graduate | 27723 | 0.13 (0.07, 0.18) | <0.001 |
| | Some College | 30870 | 0.14 (0.08, 0.19) | <0.001 |
| | College Degree | 42603 | 0.06 (0.01, 0.11) | 0.03 |
| | Graduate Degree | 15387 | −0.01 (−0.07, 0.04) | 0.65 |
| **Household income ($)** | Under 25K | 26632 | Reference | |
| | 25k to under 50k | 32049 | 0.06 (0.03, 0.09) | <0.001 |
| | 50K to under 100K | 38873 | −0.01 (−0.04, 0.02) | 0.55 |
| | 100K and over | 23229 | −0.01 (−0.04, 0.03) | 0.77 |
| **Employment** | Not full-time | 72969 | Reference | |
| | Full-time | 47814 | 0.10 (0.07, 0.12) | <0.001 |
| **Race** | Asian American | 4144 | Reference | |
| | Black | 15835 | 0.21 (0.16, 0.27) | <0.001 |
| | Hispanic | 13128 | 0.17 (0.11, 0.22) | <0.001 |
| | Other | 4795 | 0.21 (0.14, 0.28) | <0.001 |
| | White | 82881 | 0.05 (0.00, 0.10) | 0.04 |
| **Urbanicity** | Rural | 20918 | Reference | |
| | Suburban | 68710 | −0.07 (−0.09, −0.04) | <0.001 |
| | Urban | 31155 | −0.04 (−0.07, −0.01) | 0.02 |
| **At least moderate depression** | Absent | 89556 | Reference | |
| | Present | 31227 | 0.27 (0.24, 0.29) | <0.001 |
| **Political affiliation** | Democrat | 43702 | Reference | |
| | Independent/Other | 44406 | 0.20 (0.18, 0.22) | <0.001 |
| | Republican | 32675 | 0.61 (0.58, 0.63) | <0.001 |

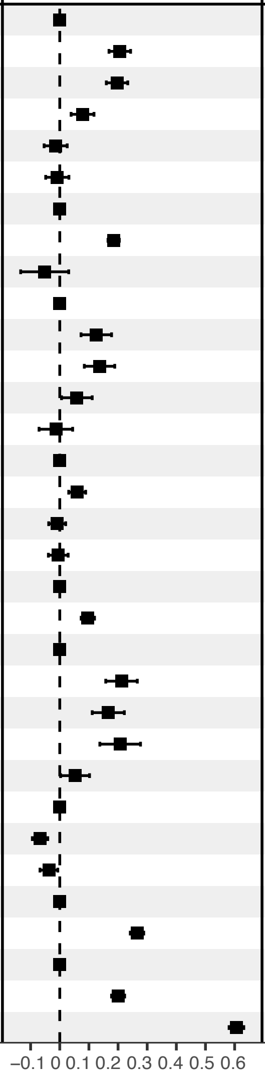

**Figure S2.** Survey-weighted logistic regression model of association between number of conspiracy items endorsed and likelihood of influenza vaccination

| Variable | | N | Odds ratio | | p |
|---|---|---|---|---|---|
| **Conspiracy items endorsed** | 0 | 21683 | | Reference | |
| | 1 | 22891 | | 1.01 (0.96, 1.06) | 0.74 |
| | 2 | 23871 | | 0.81 (0.77, 0.85) | <0.001 |
| | 3 | 14490 | | 0.66 (0.63, 0.70) | <0.001 |
| | 4 | 19137 | | 0.58 (0.55, 0.61) | <0.001 |
| **Age (years)** | 18 to 24 | 10709 | | Reference | |
| | 25 to 34 | 18578 | | 0.72 (0.67, 0.76) | <0.001 |
| | 35 to 44 | 21010 | | 0.71 (0.67, 0.75) | <0.001 |
| | 45 to 54 | 16494 | | 0.82 (0.77, 0.88) | <0.001 |
| | 55 to 64 | 15319 | | 1.31 (1.22, 1.39) | <0.001 |
| | 65 and over | 19962 | | 3.07 (2.86, 3.28) | <0.001 |
| **Gender** | Female | 61889 | | Reference | |
| | Male | 39030 | | 0.97 (0.94, 1.01) | 0.10 |
| | Nonbinary | 1153 | | 1.32 (1.14, 1.54) | <0.001 |
| **Education** | Some High School or Less | 3596 | | Reference | |
| | High School Graduate | 23539 | | 1.11 (1.02, 1.21) | 0.02 |
| | Some College | 26211 | | 1.22 (1.12, 1.33) | <0.001 |
| | College Degree | 36087 | | 1.70 (1.56, 1.85) | <0.001 |
| | Graduate Degree | 12639 | | 2.40 (2.18, 2.64) | <0.001 |
| **Household income ($)** | Under 25K | 22643 | | Reference | |
| | 25k to under 50k | 27306 | | 1.06 (1.01, 1.11) | 0.02 |
| | 50K to under 100K | 32963 | | 1.19 (1.13, 1.25) | <0.001 |
| | 100K and over | 19160 | | 1.44 (1.36, 1.52) | <0.001 |
| **Employment** | Not full-time | 62119 | | Reference | |
| | Full-time | 39953 | | 1.14 (1.09, 1.18) | <0.001 |
| **Race and ethnicity** | Asian American | 3538 | | Reference | |
| | Black | 13364 | | 0.84 (0.77, 0.92) | <0.001 |
| | Hispanic | 10910 | | 0.81 (0.74, 0.89) | <0.001 |
| | Other | 4010 | | 0.60 (0.53, 0.67) | <0.001 |
| | White | 70250 | | 0.77 (0.71, 0.84) | <0.001 |
| **Urbanicity** | Rural | 17931 | | Reference | |
| | Suburban | 58457 | | 1.20 (1.15, 1.26) | <0.001 |
| | Urban | 25684 | | 1.34 (1.27, 1.41) | <0.001 |

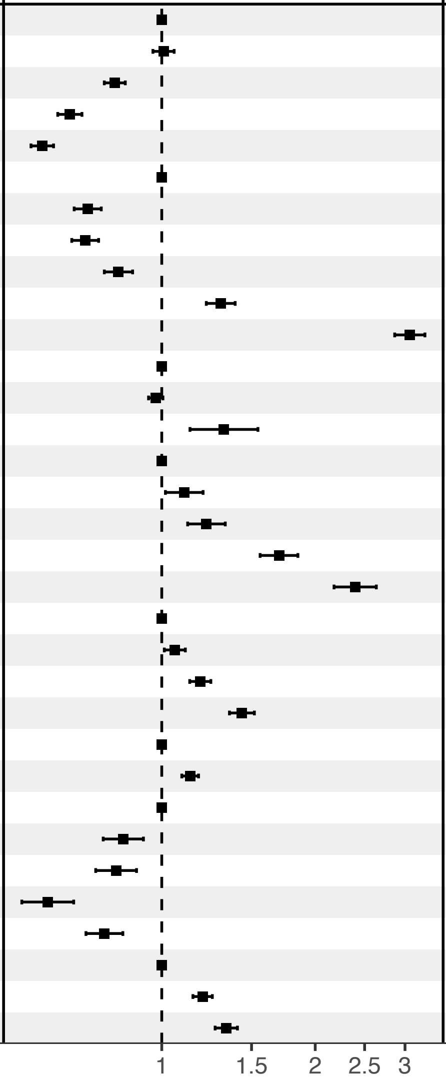

**Figure S3.** Survey-weighted logistic regression model of association between number of conspiracy items endorsed and likelihood of additional SARS-CoV-2 vaccination at subsequent survey completion

| Variable | | N | Odds ratio | p |
|---|---|---|---|---|
| **Prior number of vaccinations** | | 9059 | 0.91 (0.86, 0.95) | <0.001 |
| **Conspiracy items endorsed** | 0 | 2469 | Reference | |
| | 1 | 2122 | 1.02 (0.82, 1.27) | 0.842 |
| | 2 | 1902 | 0.91 (0.71, 1.15) | 0.429 |
| | 3 | 1015 | 0.87 (0.64, 1.18) | 0.368 |
| | 4 | 1551 | 0.72 (0.54, 0.96) | 0.023 |
| **Age (years)** | 18 to 24 | 161 | Reference | |
| | 25 to 34 | 640 | 1.77 (0.92, 3.39) | 0.086 |
| | 35 to 44 | 1052 | 1.88 (1.02, 3.50) | 0.045 |
| | 45 to 54 | 1333 | 1.84 (0.98, 3.44) | 0.057 |
| | 55 to 64 | 1916 | 1.47 (0.79, 2.74) | 0.222 |
| | 65 and over | 3957 | 2.35 (1.28, 4.34) | 0.006 |
| **Gender** | Female | 5266 | Reference | |
| | Male | 3760 | 1.00 (0.84, 1.18) | 0.976 |
| | Nonbinary | 33 | 2.27 (0.75, 6.90) | 0.149 |
| **Education** | Some High School or Less | 157 | Reference | |
| | High School Graduate | 1907 | 0.87 (0.49, 1.55) | 0.641 |
| | Some College | 2203 | 0.73 (0.41, 1.30) | 0.288 |
| | College Degree | 3580 | 0.92 (0.52, 1.62) | 0.769 |
| | Graduate Degree | 1212 | 0.76 (0.41, 1.38) | 0.361 |
| **Household income ($)** | Under 25K | 1898 | Reference | |
| | 25k to under 50k | 2731 | 0.95 (0.74, 1.22) | 0.680 |
| | 50K to under 100K | 3003 | 1.13 (0.88, 1.46) | 0.340 |
| | 100K and over | 1427 | 1.23 (0.92, 1.66) | 0.166 |
| **Employment** | Not full-time | 6847 | Reference | |
| | Full-time | 2212 | 1.16 (0.94, 1.43) | 0.175 |
| **Race and ethnicity** | Asian American | 520 | Reference | |
| | Black | 933 | 1.15 (0.79, 1.67) | 0.469 |
| | Hispanic | 475 | 0.93 (0.60, 1.46) | 0.759 |
| | Other | 216 | 0.74 (0.40, 1.38) | 0.346 |
| | White | 6915 | 0.87 (0.63, 1.20) | 0.405 |
| **Urbanicity** | Rural | 1600 | Reference | |
| | Suburban | 5367 | 1.14 (0.90, 1.44) | 0.265 |
| | Urban | 2092 | 1.09 (0.83, 1.43) | 0.543 |

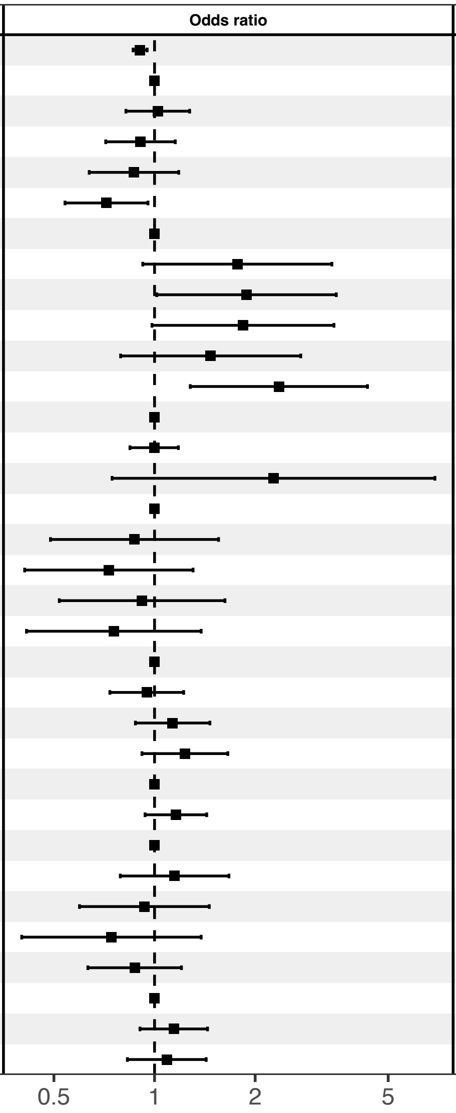
